# Business as Un-usual: Access to mental health and primary care services for people with severe mental illness during the COVID-19 restrictions

**DOI:** 10.1101/2021.05.24.21257694

**Authors:** Elizabeth Newbronner, Panagiotis Spanakis, Ruth Wadman, Suzanne Crossland, Paul Heron, Gordon Johnston, Lauren Walker, Simon Gilbody, Emily Peckham

## Abstract

**Aims:** To explore: how satisfied people with severe mental illness (SMI) are with the support received during the pandemic; understand any difficulties encountered when accessing both mental health and primary care services; consider ways to mitigate these difficulties; and assess the perceived need for future support from mental health services.

**Materials and Methods:** A representative sample was drawn from a large transdiagnostic clinical cohort of people with SMI, which was recruited between April 2016 and March 2020. The sample was re-surveyed a few months after the beginning of the restrictions. Descriptive frequency statistics were used to analyse the quantitative data. The free text responses were analysed thematically.

**Results:** 367 participants responded to the survey. Two thirds were receiving support from mental health services with the rest supported in primary care or self-managing. A quarter thought they would need more mental health support in the coming year. Half had needed to used community mental health services during the pandemic and the majority had been able to get support. A minority reported that their mental health had deteriorated but they had either not got the supported they wanted or had not sought help. The biggest service change was the reduction in face-to-face appointments and increasing use of phone and video call support. Nearly half of those using mental health services found this change acceptable or even preferred it; acceptability was influenced by several factors. Participants were more likely to be satisfied with support received when seen in person.

**Discussion:** Although most participants were satisfied with the mental health support they had received, a minority were not. This, couple with findings on future need for mental health support has implications for post pandemic demand on services. Remote care has brought benefits but also risks that it could increase inequalities in access to services.

## Introduction

In mid-March 2020, soon after the WHO declared the outbreak of COVID-19 to be a pandemic, sweeping changes in the way mental health and primary care services in the UK are provided were introduced. A survey of mental health staff, conducted early in the first wave of the pandemic in the UK (i.e April/May 2020) found that the number of face-to-face meetings was reduced, and phone and video appointments and support were rapidly introduced [1]. This observation was supported by a recent report from the House of Lords Covid-19 Committee [2]. In most areas of the UK the frequency of contacts and the range of both NHS and third sector services available also reduced [3]. Research is beginning to explore the impact of the pandemic and the pandemic restrictions on people with pre-existing mental health conditions. However, there has been little research into the specific experiences of people living with severe mental illness (SMI), and in particular how their access to mental health and primary care services may have changed.

People with SMI are an especially vulnerable group who already experience significant health inequalities. Notably they currently experience a mortality gap of 15-20 years when compared to people without SMI [4, 5, 6]. An important driver for this gap is preventable physical health conditions, linked to behavioural risk factors, such as poor diet and smoking. However, access to and take-up of services is also a factor [7, 8]. There are concerns that the changes in the way services have been delivered during the pandemic may have further increased the barriers to access that people with SMI experience (e.g. access to digital technologies or familiarity/confidence in using them) [9,10]. It is therefore important to: explore how satisfied people with SMI are with the support they received during the pandemic; understand any difficulties they encountered when accessing both mental health and primary care services; consider ways to mitigate these difficulties; and assess the perceived need for future support from mental health services. The aim of this study was to use a large clinical cohort of people with SMI, which was recruited in the years immediately prior to the pandemic restrictions and was re-surveyed a few months after the beginning of the restrictions, to explore these issues.

## Materials and Methods

The Closing the Gap (CtG) study is a large (n=9,914) transdiagnostic clinical cohort recruited between April 2016 and March 2020. Participants have documented diagnoses of schizophrenia or delusional/psychotic illness (ICD 10 F20.X & F22.X or DSM equivalent) or bipolar disorder (ICD F31.X or DSM-equivalent). The composition of the CtG cohort has previously been described [11].

We were funded to explore the impact of the COVID-19 pandemic in a sub-section of the CtG clinical cohort and we identified participants for Optimising Well-being in Self-Isolation study (OWLS) (https://sites.google.com/york.ac.uk/owls-study/home). To ensure that the OWLS COVID-19 sub-cohort was representative we created a sampling framework based on gender, age, ethnicity and whether they were recruited via primary or secondary care. OWLS participants were recruited from 17 mental health trusts and six primary care Clinical Research Networks (across urban and rural settings in England).

To be eligible to take part in OWLS COVID-19 study, people had to be aged 18 or over, have taken part in the CtG study and have consented to be contacted again about further research. They must also have been originally recruited from a clinical site with capacity to support the OWLS study. This approach enabled us to create longitudinal data linkage and to rapidly identify participants during the COVID-19 pandemic. To increase the chances of participants responding and their contact details remaining current, we invited the participants who had been more recently recruited to the CtG cohort. People who met the eligibility criteria were contacted by telephone or letter and invited to take part in the OWLS COVID-19 study. Those who agreed to take part were offered three options; i) complete the survey over the phone with a researcher, ii) be sent a link to complete the survey online or iii) be sent a hard copy of the questionnaire in the post to complete and return.

In Section 2 of the survey, we explored how participants’ access to mental health and primary care services had changed during the pandemic. For example, we asked participants who were receiving support from mental health services, whether they had experienced one or more specific change (e.g. seeing a different mental health worker to the person they normally saw). If they had, they could indicate how they felt about the change with one of three answers (i.e. I like it better; its ok – not better or worse; or I don’t like it). The full OWLS 1 questionnaire is shown in the supplementary materials (S1). For all services, we were interested to understand how changes in the way services were provided, in particular the shift to phone and online meetings and appointments, might have affected peoples’ experiences. We also wished to explore what factors might have hindered or facilitated good care and support, despite the change in access to services. For mental health services we also asked people about the support they anticipated needing from services in the year ahead, an issue of particular interest to NHS providers.

The study analysis plan was registered on Open Science Framework (available at https://doi.org/10.17605/OSF.IO/E3KDM - section 2.1). Descriptive frequency statistics (Ns and percentages) were used to describe key sample characteristics and service use variables. 2×2 cross-tabulations and chi-square tests were used to examine the association between a) unmet need for support and changes in mental or physical health, and b) mode of service delivery and service satisfaction. The latter was examined for each service type separately (GPs, Community mental health, Mental health crisis services) and p values were corrected for multiple testing (p multiplied by four). For brevity, the results for general hospital services are not included in the main text of the paper but are available in the supplementary materials (S2). The statistical significance criterion used for all analyses was p < 0.05. Analyses were undertaken using SPSS v.26.

In the pre-registered analysis plan it was stated that mode of service delivery would be coded at three-levels (in person, over the phone, or online e.g. video call). However, too few participants reported receiving services online (N ranged from 2 to 14, depending on service type) to allow for meaningful comparisons, and so this was merged with receiving services over the phone into a remote delivery category. The plan also stated that association between Community mental health service satisfaction and mode of service delivery would be stratified per primary or secondary care patients. As very few participants currently in primary care reported getting support from Community mental health services (N = 24) to allow for meaningful comparisons, analysis was conducted only in the full sample.

At the end of the survey there was a free text box where respondents could add comments about any aspect of their experiences during the pandemic. 147 (40%) respondents chose to do so. These free text responses we transferred to NVivo and then analysed thematically [12]. The seven sections of the survey provided the initial structure for the analysis. Fifty-two respondents added comments which in some way related to their access to or experience of using of services. Quotations from the free text analysis are used in this paper to illustrate or bring to life the findings from the quantitative data.

The authors assert that all procedures contributing to this work comply with the ethical standards of the relevant national and institutional committees on human experimentation and with the Helsinki Declaration of 1975, as revised in 2008. All procedures involving human subjects/patients were approved by the North West – Liverpool Central Research Ethics Committee (reference 20/NW/0276). Written or verbal informed consent was obtained from all subjects/patients.

## Results

In this section we begin by briefly describing the demographic characteristics of the OWLS participants. We then present our findings in relation to participants experiences of accessing both community mental health services and mental health crisis services, and their perceived need for future support from mental health services. We then go on to describe their experiences of using primary care services.

The first survey for the OWLS COVID-19 study, which was conducted between July to December 2020, recruited 367 people from the CtG Cohort. Table 1 describes the socio-demographic characteristics and diagnosis of OWLS participants. It should be noted that it was an ethical requirement of the CtG study that the participants consent to their diagnosis being provided to the research team and some participants did not consent to this. In the interests of inclusivity and because there may be some differences between those who consented to their diagnosis being provided and those who did not, we did not make it a requirement of OWLS that the participants should consent to their diagnosis being provided. The mean age was 50.5 (range = 20 to 86, SD ± 15.69) with 51.0% male and 77.4% white British.

**Table 1:**
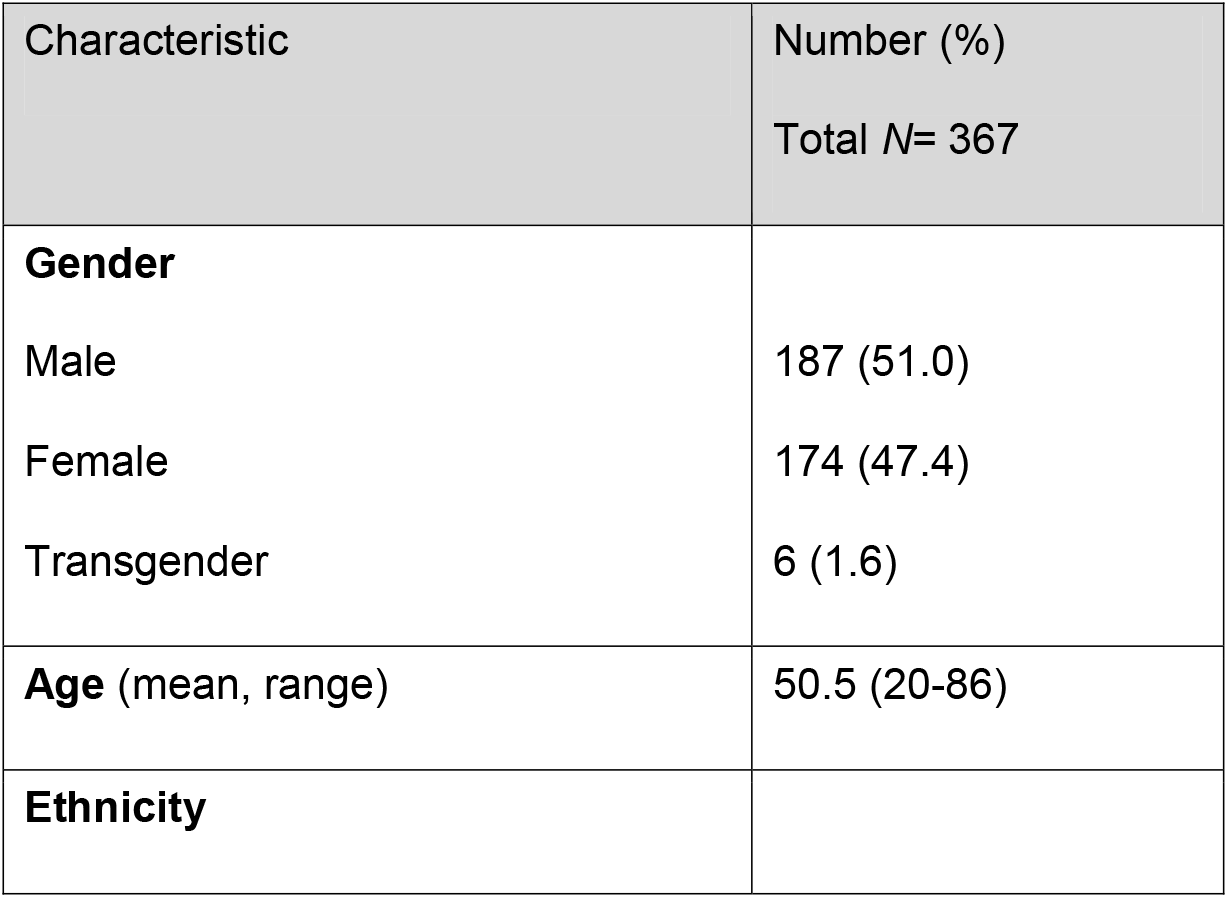
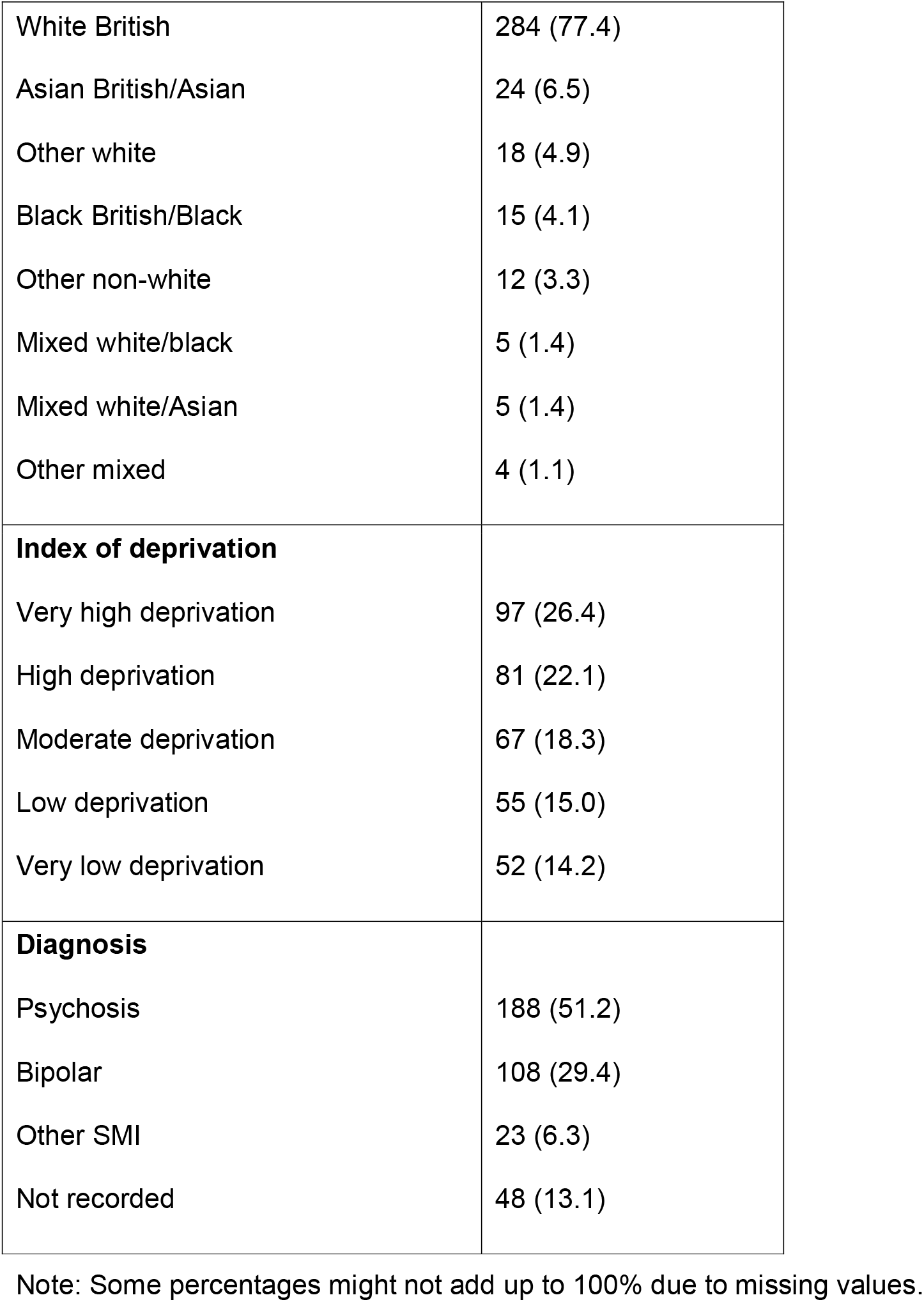
Demographic characteristics of OWLS COVID-19 study SMI population during pandemic restrictions.

## Mental Health Services

Of the 367 people who completed the survey, 224 (61%) were receiving support from mental health services, with the rest being supported in primary care or self-managing.

### Community Mental Health Services

Just over half of the survey respondents (194/52.9%) had needed to used community mental health services during the pandemic and the overwhelming majority reported that they had been able to get support. The quotation below highlights the creative approaches sometimes adopted by mental health staff to maintain support:

> *“My CPN* [Community Psychiatric Nurse] *was very supportive and has organised my Clozaril bloods to be taken in my garden so I don’t need to travel all the way to [TOWN]. I receive regular telephone calls from my CPN and face to face visits every 2 to 3 weeks*.*”*

We examined whether there was any difference between those who said that their mental health had deteriorated and those who said it had not, in terms of getting support. As Table 2 below shows there was no significant association between deterioration in mental health and receiving mental health services (χ^2^(1) = .99, p = .319). However, 14.5% (n=12) did report that their mental health had deteriorated but they had either not got the supported they wanted or had not sought help. One of the free text comments illustrates this: *“Before the pandemic I wasn’t using MH services but as my MH has gotten worse then I have become very conscious of not being able to access services”*.

**Table 2.** Change in mental health and support from community mental health services.

| Mental health change since beginning of pandemic | Did you get support from community mental health services? |  |  |
| --- | --- | --- | --- |
|  | Yes (N = 164) | No/Haven't tried (N = 22) | Total |
| Deterioration | 71 (85.5%) | 12 (14.5%) | 83 |
| No deterioration | 93 (90.3%) | 10 (9.7%) | 103 |

Note: 8 missing. Percentages are per row.

Our Lived Experience Advisory Panel suggested that for many people with mental health needs, having confidence that support would be forthcoming if required, was important for peoples’ wellbeing. So, we asked all survey respondents how confident they felt about support being available, should they need it. More than half (207/56.4%) felt they would be able to access help, but a substantial minority (158/43.1%) were not confident that support would be available.

### Changes to the Way Mental Health Services were Provided

The pandemic led to substantial and rapid changes in the way in which mental health services were provided. The biggest shift was the reduction in face-to-face appointments and increasing use of phone and online consultations and support. However, people also experienced less frequent contact with services, or had a more limited range of services (including community and voluntary sector services) available to them. A few people saw a different mental health worker. We asked the people who were currently receiving support from mental health services (n=224) how they felt about these changes and 221 provided responses. Table 3 provides and overview of their responses.

**Table 3.** Views about change in the way mental health services were provided.

|  | I have not<br>experienced<br>this change | I like it<br>better | Its OK – not<br>better or<br>worse | I don't like it |
| --- | --- | --- | --- | --- |
| I had support on the<br>telephone/online instead of<br>face-to-face appointments | 44 (19.9%) | 32 (14.5%) | 74 (33.5%) | 71 (32.1%) |
| I had less frequent contact<br>with mental health services | 105 (47%) | 12 (5.4%) | 59 (26.7%) | 45 (20.4%) |
| I had more frequent contact<br>with mental health services | 147 (67%) | 19 (8.7%) | 41 (18.8%) | 11 (5%) |
| I had a more limited range of services or support | 104 (47.7%) | 9 (4.1%) | 45 (20.6%) | 60 (27.5%) |
| I saw a different mental health worker to the person who would normally support me | 149 (68.7%) | 9 (4.1%) | 40 (18.4) | 19 (8.8%) |
| I went to a different place for appointments or support | 170 (79.1%) | 8 (3.7%) | 22 (10.2%) | 15 (7%) |

Note: Percentages are per row

The results show that despite the impact of the pandemic, a high proportion of those survey respondents who were in contact with mental health services, were seeing the same mental health worker, often in the same place. Clearly many people were being supported on the telephone or online instead of face to face, and whilst almost a third did not like this change, nearly half found it acceptable or even preferred it.

The findings from the OWLS study in relation to access to digital devices are reported in a separate paper. However, it is useful to note here that the majority of study participants did have access to a smartphone, tablet, laptop and/or desktop computer. However, 13.4% (n=49) did not have any of these devices. The free text comments also suggested that even where people did have access to such devices, they may not have exclusive use (e.g. a ‘family’ computer), their internet access may be limited or there may be practical and emotional concerns associated with online appointments.

Focusing specifically on the 168 people who had received support from community mental health services, just over half (n=89/52.7%) had received that support in person. Sixty-six people (39.1%) had support over the phone and a minority (n=14/3.8) had some kind of online video support (e.g. NHS Attend Anywhere). Almost two thirds (n=112/65.9%) were completely satisfied with the support they received but as Table 4 below shows, there was a significant association between satisfaction with the support received from community mental health services and the way in which it was provided (χ^2^(1) = 22.92, p < .001). Specifically, the proportion of those who reported receiving support in person and were completely satisfied was significantly higher that the proportion of those who reported receiving support remotely and were completely satisfied.

**Table 4.** Satisfaction with support from community mental health services.

| Did you receive the level of support you needed from your Community Mental Health Services? | How was your appointment delivered? |  |
| --- | --- | --- |
|  | In person (N = 89) | Phone/Online (N = 79) |
| Completely | 73 (82.0%) | 37 (46.8%) |
| Partly/No | 16 (18.0%) | 42 (53.2%) |

Note: Percentages are per column

Many of the free text comments added by respondents were concerned with the shift to phone and online support, including practical difficulties this created:

> *“I have had a referral back to mental health services, and I am due to have a phone appointment with a doctor. I will have to find a quiet place to go during work time which will not be easy and I’m worried that I might be overhead. The last appointment was three hours later than scheduled so I am also worried that it will conflict with other appointments or meetings I have that day*.*”*

People also highlighted the importance of having an established relationship with their mental health professional: *“Fortunately, I got to know my new CPN through a few face-to-face visits and built a rapport so that now that we speak online it is okay - without the face-to-face it would have been more difficult”*. Others emphasized how much they valued face to face contact:

> *“I have had 3 appointments with my psychiatrist over the phone during the lockdown and found this to be a poor substitute compared to a face-to-face meetings. At first my CPN called me once a week for about a 20 minute phone call then we used Attend Anywhere the NHS video calling service. Finally, for the last month of the previous lockdown we met in the garden of the local CMHT under a gazebo wearing masks and social distancing. It was great to see my CPN again after so many months and gave me a real boost*.*”*

Interestingly, a small number of people said they preferred the option of phone and online support, finding it more convenient or less stressful: *“I don’t need to be stressed about going out or on the bus, things that make my paranoia and voices worse. And I don’t have to leave my dog on her own”*.

### Mental Health Crisis Services

Almost a fifth of respondents (n=59/16.1%) had needed to use mental health crisis services. Over three quarters of this group (n=45/76.3%) were able to get support but a significant minority (n=14/23.7%) reported that they either did not get the support they needed or did not try to get help. The free text comments suggest that for this group the implications of not being able to access support were potentially very serious:

> *“Just before lockdown I was trying to manage without medication. I then hit crisis point over a bank holiday period during lockdown. After several calls to the crisis team I was unable to get any help. It wasn’t until a suicide attempt did I start to get help. I have only asked for help twice in seven years, so have never abused the service. The overstretched system was sadly lacking in an extreme time of need*.*”*

We also explored satisfaction with the way in which support was provided (see Table 5). We found there was no significant association between receiving the level of support needed from mental health crisis services and the way in which this support was provided (χ^2^(1) =.07, p = 4.000) (i.e. the proportion of those who were completely satisfied with the support they received was broadly similar for the in person and phone/online groups).

**Table 5.** Satisfaction with support from mental health crisis services.

| Did you receive the level of support<br>you needed from you Mental Health<br>Crisis Services? | How was your appointment delivered? |  |
| --- | --- | --- |
| | In person ($N = 20$) | Phone/Online ($N = 25$) |
| Completely | 12 (60.0%) | 14 (56.0%) |
| Partly/No | 8 (40.0%) | 11 (44.0%) |

Note: Percentages are per column.

### Future Need for Mental Health Services

There is substantial interest within the NHS about the impact of the COVID-19 pandemic on future need for mental health services and we were able to explore this to a limited extent in this study. We asked those people who were not currently getting support from mental health services whether they thought they would need support in the year ahead, and just under a third (n=44/31.7%) thought they would.

For those people who were already being supported by mental health services, we asked how their need for support might change in the year ahead. As Table 6 below shows, just over a quarter thought they would need more support either because their mental health had declined, or they had been putting off dealing with some issues, or they felt the support they had prior to the pandemic was insufficient, and almost a quarter thought they might need more support.

**Table 6.** Perceived need for support from mental health services in the year ahead.

| About the year ahead, which of the following fits you best? | *N* (%) – out of total *N* = 224 |
| --- | --- |
| I think I will need the same or less support – my mental health has improved during the pandemic | 37 (16.5) |
| I don't think the support I need will change – I've been ok during the pandemic. | 71 (31.7) |
| I might need more support – the pandemic has been difficult for me but I am going to see how things go. | 54 (24.1) |
| I will need more support – during the pandemic my mental health has got worse | 32 (14.3) |
| I will need more support – I put off dealing with some things during the pandemic but now I want to get advice/help with them. | 18 (8.0) |
| I didn't have enough support before the pandemic and I still need more support | 10 (4.5) |

Note: This question was only asked of participants who were currently getting support from mental health services.

### Primary Care Services

Two thirds of the survey respondents (n=240) had needed to use their GP practice services during the pandemic (for mental and/or physical health conditions). Of this group, the majority (n=207/86%) reported that they had been able to get an appointment and most (n=154/74.4%) felt they had received the care and support they needed. However, like mental health services, GP practices also moved to much greater use of phone and video consultations. We examined whether there was any association between deterioration in physical health and access to GP care and support. Table 7 below shows that there was no significant association (χ^2^(1) = .14, p = .706).

**Table 7.** Changes in physical health and support from GP services.

| Physical health change since beginning of pandemic | Did you get support from GP services? |  |  |
| --- | --- | --- | --- |
| | Yes ($N = 202$) | No/Haven't tried ($N = 29$) | Total |
| Deterioration | 77 (38.1%) | 10 (34.5%) | 87 |
| No deterioration | 125 (61.9%) | 19 (65.5%) | 144 |

Note: Percentages are per row.

We then examined whether people felt they had received the support they needed. Almost three quarters reported that they had but there was a marked different between those who had had a face-to-face appointment and those supported on the phone or online. As Table 8 shows, people were more likely to feel completely satisfied with the support they received when it was face-to-face (χ^2^(1) = 10.46, p = .008).

**Table 8.**
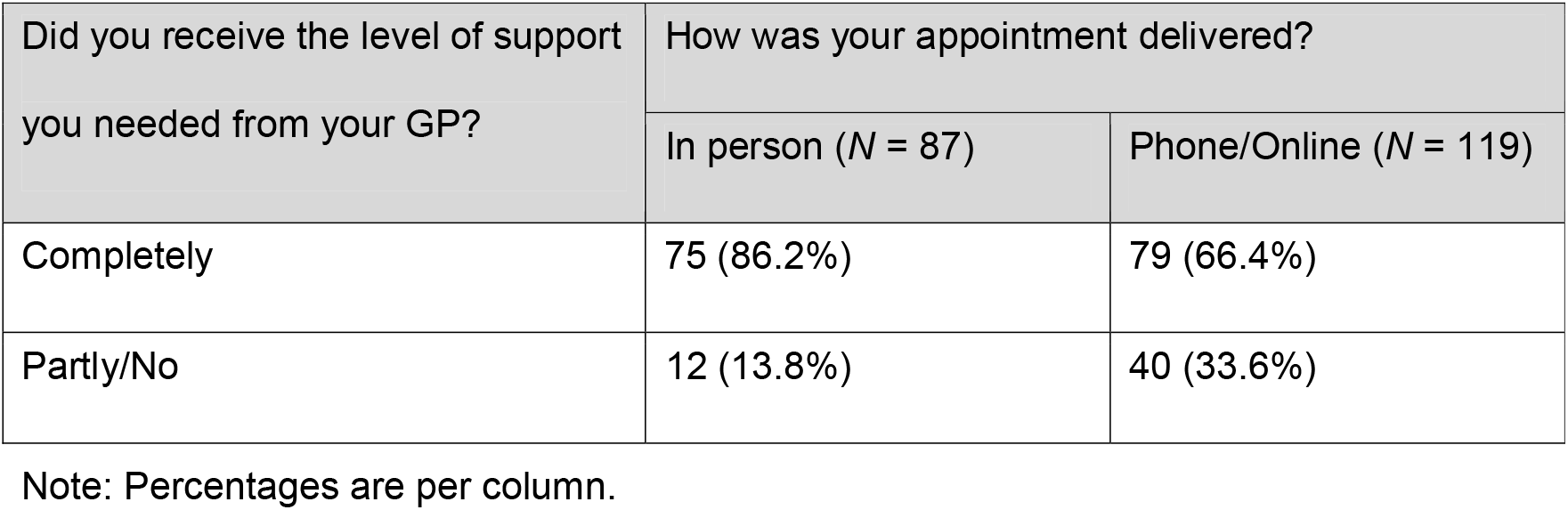
Satisfaction with support from GP services.

In relation to medication and pharmacy services, most survey respondents (n=330/89%) had needed prescription medication during the pandemic and almost all were able to obtain their medication from their usual pharmacy. However, the free text comments revealed that a few people had experienced delays and difficulties with the supply of their usual medication. One respondent said that they were on three types of medication, and at one point had to go to three different pharmacies to get the medication they needed. They noted that they were relatively well but felt that if they had not been, they might have stopped taking their medication. Another explained:

> *“There was a shortage of medication supply in the area. I struggled to get my prescription tablets and when I eventually got them then I got a slightly different type (tablets vs capsules). I wasn’t sure what would replace them and how I would deal with them - at the time I thought they were going to give me a different drug and the ones I take are quite a fine balance. So that was very worrying*.*”*

## Discussion

Most participants reported that they had been able to access support from community mental health services and a relatively high proportion said they were satisfied with the support they received. This contrasts with the findings from a qualitative study by Gillard et al [10] in which participants reported issues around continuity of care, not getting treatment as usual and service changes. However, in our study a minority did report that their mental health had deteriorated but they had either not got the supported they wanted or had not sought help. This is an issue of concern, which alongside our findings on future need for increased support, may have implications in terms of post-pandemic increased demand for mental health services. Furthermore, for some people living with severe mental illness, the fear of becoming unwell and not being able to access support is a source of anxiety. A substantial minority of participants in our study did not feel confident that support would be available should they need – a perception that mental health services may need to explore and address.

The biggest change in the way in which mental health services were provided was the reduction in face-to-face appointments and the increase in remote care. We found that almost half those who had used community mental health service had received support in person, with staff using creative approaches to maintaining face-to-face contact. Many people were being supported on the telephone and a few had online support. Whilst almost a third did not like this change, nearly half found it acceptable or even preferred it. However, the free text comments suggest that acceptability was influenced by several factors. In particular, respondents felt that telephone or video appointments were far easier when: they had an established relationship with their mental health professional; where they had somewhere private to hold a telephone conversation; or they had access to a (private) digital device - findings which echo those of a recent qualitative study by Liberati et al [13]. Furthermore, whilst increased use of remote mental health care might be acceptable for a limited period, during a public health crisis, in the long term it could lead to problems with accessibility, equity of care and service quality [9,10,13]. The limited availability of online support reported by participants in our study, suggests that Greenhalgh et al [14] are correct in suggesting that the evidence relating to video-based remote care needs to be strengthened. Interestingly, whilst people were more likely to be satisfied with the support they received from community mental health service when they had been seen in person, in mental health crisis services the way support was provided appeared less important. We speculate that this could be because phone support is already commonly used in crisis services and/or because remote care facilitated more rapid access to support.

With regard to access to primary care services, the majority of participants had been able to get an appointment and most felt they had received the care and support they needed. However, those who had a face-to-face appointment were more likely to feel satisfied with the support they received. Interestingly, the free text comments revealed a specific issue in relation to obtaining medication, with a number of respondents reporting delays and difficulties with the supply of their usual medication.

Overall then, our study found that the majority of people with SMI were able to access support but those who received remote support were less likely to be satisfied with that support. Furthermore, the context of remote support was important. This suggests that if, as seems likely, service providers continue to employ some element of remote care, service users should have a choice about whether they want remote care and the mode of remote support they receive. More broadly, as services continue to adapt to the prevailing pandemic situation and review future provision, there should be a renewed focus on the implications of intersectionality. Specifically, how the experience of living with severe mental illness, coupled with other vulnerabilities or disadvantages such as disabling physical conditions, low income, and ethnicity can create even greater inequalities in access to services.

The findings from this study suggest that some people with SMI do not feel that they have received enough support during the pandemic, or have been reluctant to seek support, perhaps because of the way services were provided or fear of contracting the virus. As a consequence, when the pandemic restrictions ease there could be an increase in demand from mental health services from existing service users. However, this may be tempered by the fact that many mental health professionals and GPs appear to have prioritised support for people with SMI and found creative ways to maintain some face-to-face contact.

Some of the changes introduced as a result of the COVID-19 pandemic will almost certainly continue to be part of future mental health and primary care service. These changes can bring benefits for service providers and some service users. However, they could also increase the risk of unequal access to support and poorer quality of care. To counter these risks, people with SMI must be consulted about the nature of any long-term changes, and going forward, close attention needs to be paid to services users’ individual circumstance and preferences.

## Data Availability

All key data are within the manuscript and its Supporting Information files. Additional
information is available on request from the corresponding author.

## Acknowledgements

We would like to thank all the participants in the OWLS study, the lived experience panel who provided advice throughout the study, and the mental health trust and CRN staff who supported study recruitment.

## Supporting Information

S1 OWLS 1 Survey

S2 Results for General Hospital Services

